# Lifestyles according to disease duration in patients with diabetes and hypertension

**DOI:** 10.1101/2024.08.17.24311880

**Authors:** Joan A. Loayza-Castro, Lupita Ana Maria Valladolid-Sandoval, Luisa Erika Milagros Vásquez Romero, Fiorella E. Zuzunaga-Montoya, Jhonatan Roberto Astucuri Hidalgo, Víctor Juan Vera-Ponce

## Abstract

**Introduction:** In patients with hypertension (HTN) and diabetes mellitus (DM), lifestyles should be optimal to avoid long-term complications. However, it is sometimes unclear whether the disease duration factor could contribute to their improvement.

**Objective:** 1) To analyze the trend of lifestyles in patients with DM or HTN; 2) to determine the association of lifestyles according to the time since diagnosis in these patients.

**Materials and methods:** Analytical cross-sectional study based on Peru’s Demographic and Family Health Survey between 2014 and 2023. Lifestyles were: smoking status, alcohol consumption, and fruit/vegetable consumption. The time since diagnosis variable was dichotomized (< 2 years versus ≥ 2 years).

**Results:** Lifestyle trends between the established years showed a fluctuating pattern, with a significant interruption in 2020, followed by a return to pre-2020 variations, except for a decrease in smokers in subjects with HTN. On the other hand, no significant association was found between any of the three lifestyles and the time since diagnosis of HTN or DM2.

**Conclusion:** The trends found in lifestyles have not resulted in a sustained decrease in bad habits or a constant improvement in healthy lifestyles, except for smokers in patients with HTN, but rather a general state without substantial changes has been maintained. In turn, the disease duration does not seem to be a determining factor for changes in the lifestyles of patients with DM or HTN.

## Introduction

Chronic non-communicable diseases are currently one of the main problems in global public health, among which Diabetes and Hypertension can be highlighted due to their high prevalence^(1)^. Globally, it is estimated that the prevalence of diabetes is approximately 9.3%, and in relation to arterial hypertension, it is approximately 31% of the adult population ^(2,3)^. In Latin America, these values are not different, as the prevalence of diabetes varies between 8% and 13%, and the prevalence of HTN reaches up to 30% in some populations ^(4,5)^. In Peru, a prevalence of diabetes close to 7.3% and hypertension of 24.9% in the adult population has been reported ^(6,7)^.

Considering the high prevalence and impact of diabetes and hypertension on public health, it is important to explore factors that could influence the progression and management of these diseases, such as optimal lifestyles, which play an interesting role in reducing morbidity and mortality from these diseases ^(8)^.

Lifestyles play an important role in the management of diabetes and hypertension. Factors such as diet, physical exercise, smoking, and alcohol consumption have a direct association with the control of these chronic diseases. Various studies have shown that lifestyle interventions are not only effective for the prevention of these diseases but also for improving management and reducing the incidence of long-term complications ^(9)^. However, it is important to recognize that these habits and behaviors can evolve over time, depending on the duration of the disease.

The time since diagnosis is a key factor to consider in the changes and adaptations that patients experience in their lifestyle. As they live with these diseases for prolonged periods, they tend to adopt healthier behaviors as part of their management strategy. However, evidence suggests that those with a recent diagnosis may face greater difficulties in developing these healthy habits ^(10)^.

Thus, this study presents the directed hypothesis that lifestyles are different according to the duration of the disease, and that it could even be better with more patient experience with their pathology. For this reason, the objective of this study was: 1) to analyze the trend of lifestyles in patients with DM or HTN during the period 2014 - 2023; 2) to determine the association of lifestyles according to the time since diagnosis in these patients.

## Methods

### Study Design and Context

This is an analytical cross-sectional study based on data from the Demographic and Family Health Survey (DHS) of Peru between 2014 and 2023 ^(11)^. DHS is a national study that aims to obtain representative information of the entire Peruvian population, differentiating between urban and rural areas, as well as among the three natural regions of the country: Coast, Highlands, and Jungle.

### Population, Eligibility Criteria, and Sample

DHS was designed for the Peruvian population of all ages, taking into account participation from rural and urban areas of all departments of Peru. The DHS sampling was based on a probabilistic, stratified, two-stage design, differentiating urban and rural areas.

For this study, only individuals aged 18 years and older who were aware of their diagnosis of hypertension and type 2 diabetes mellitus were included. Subjects who did not have data on the time since they received the diagnosis were excluded.

### Variables and Measurement

The main variables were two:

1. Lifestyles: Lifestyle was determined by evaluating the variables of smoking status, alcohol consumption, and fruit and vegetable consumption
  a. Smoking status: Obtained from the participant’s self-report, and classified as never smoked, former smoker, current smoker, and daily smoker. For the regression analysis, this variable was dichotomized into never/former smoker versus current smoker/daily smoker.
  b. Alcohol consumption: This variable was defined by the participant’s self-report and categorized as never or has not consumed alcohol in the last 12 months, non-excessive consumption (less than or equal to one occasion in the last 30 days, but less than 5 drinks for men or less than 4 drinks for women during the last 12 months) and excessive consumption (more than one occasion in the last 30 days and more than 5 drinks for men and more than 4 drinks for women) ^(12,13)^. However, for the regression analysis, it was dichotomized into never/non-excessive consumption versus excessive.
  c. Fruit and vegetable consumption: This variable was worked considering consumption greater than 5 portions per day (yes versus no).
2. Time since diagnosis: This variable was obtained from the questions asked to participants: “How long ago were you diagnosed with high blood pressure/high blood sugar?” These variables were categorized with “yes” and “no” responses.

Regarding sociodemographic variables, the following were evaluated: sex (female or male), age (categorized as 18 to 59 years and 60 years or older), educational level (no education or primary and secondary and higher), natural region (Metropolitan Lima, Rest of the coast, Highlands and Jungle), wealth index (very poor/poor, medium or rich/very rich and area of residence (urban or rural) and nutritional status (normal weight, overweight and obesity).

### Procedures

The DHS in Peru has evolved in its data collection methods over time. Until 2015, paper questionnaires were used, but digital tablets were implemented in 2016. Data collection has been carried out following internationally standardized and validated protocols, with trained personnel from the National Institute of Statistics and Informatics conducting interviews and anthropometric measurements in selected households.

For the measurement of Body Mass Index (BMI), daily calibrated anthropometric equipment is used. Weight is determined with SECA model 874 electronic scales, with a precision of 0.1 kg, while height is measured with SECA model 213 portable stadiometers, with a precision of 1 mm.

Participants are weighed in light clothing without shoes, following the Frankfurt protocol for head position. Each measurement is performed in duplicate, using the average for final calculations. In case of discrepancies greater than 0.5 kg in weight or 1 cm in height, a third measurement is taken.

Blood pressure is measured using OMRON model HEM-7130 automatic digital sphygmomanometers, clinically validated. Two cuff sizes are used to adapt to participants’ arm circumference. Measurements are taken after the participant has been seated and at rest for at least 5 minutes, with the right arm supported at heart level. Two readings are taken with a two-minute interval between them. If the difference between readings is greater than 10 mmHg for systolic or 6 mmHg for diastolic pressure, a third measurement is taken. The final value is calculated as the average of the two (or three) readings.

During the COVID-19 pandemic in 2020 and 2021, the DHS methodology had to adapt to the circumstances. Many interviews were conducted by telephone, and physical measurements were carried out after the mandatory social isolation period decreed by the Peruvian government. This adaptation allowed for the continuation of crucial data collection, albeit with some modifications in the process. Fortunately, by 2022, the face-to-face development of the survey could be carried out normally, resuming the standard procedures established before the pandemic.

### Statistical Analysis

For the statistical analysis of this research, R Studio version 4.2.1 was used, taking advantage of various functionalities and specialized packages to ensure a robust and comprehensive analysis.

First, an exhaustive descriptive analysis of all study variables was performed to generate an overview of the sociodemographic characteristics of the database. For categorical variables, absolute and relative frequencies (presented as percentages) were calculated. For continuous variables, measures of central tendency (mean or median) and dispersion (standard deviation or interquartile range) were calculated, according to the distribution of the data. For this purpose, functions from the ‘summarytools’ package in R were used.

On the other hand, trend graphs covering the study period between 2014 and 2023 were developed to evaluate variations of the variables of importance for the study over time.

A Poisson regression model with robust variance was used to calculate crude (cPR) and adjusted (aPR) prevalence ratios with their respective 95% confidence intervals (95% CI). This model allowed for estimating the association between variables of interest, controlling for possible confounding factors.

It is important to note that all analyses were performed taking into account the complex design of the DHS survey, using the provided sampling weights. To incorporate the survey design into the analyses, the ‘survey’ package in R was used, ensuring that the results are representative of the target population and that the estimates are accurate and reliable.

### Ethical Aspects

By using the DHS survey, information is freely accessible. The data do not contain any personal specifications and there was no contact with the participants. Therefore, a review by an ethics committee was not required to conduct this study. However, it should be considered that this research has followed ethical principles aligned with the Declaration of Helsinki.

## Results

Table 1 presents the characteristics of participants with hypertension (n = 11,989) and diabetes (n = 4,022). In both groups, the majority were women (62.36% and 57.39% respectively), over 60 years old (70.09% and 63.51%), with secondary or higher education (57.89% and 65.80%), residing in urban areas (76.45% and 87.53%), and belonging to the highest wealth quintiles (46.62% and 55.97%). Regarding lifestyles, similar patterns were observed in both groups: low consumption of fruits and vegetables (≥5 daily portions: 8.87% in hypertensive, 9.98% in diabetics), low prevalence of daily smokers (2.10% and 2.56%), and low excessive alcohol consumption (2.26% and 2.73%). The majority had a diagnosis of their condition for 2 years or more (61.41% in hypertension, 69.74% in diabetes).

**Table 1.** Characteristics of the study sample.

| Characteristic | Hypertension<br>n = 11,989 | Diabetes<br>n = 4,022 |
| --- | --- | --- |
| **Sex** |  |  |
| Female | 7,476 (62.36%) | 2,308 (57.39%) |
| Male | 4,512 (37.64%) | 1,714 (42.61%) |
| **Group age** |  |  |
| 18 to 59 years old | 2,589 (29.91%) | 1,104 (36.49%) |
| 60 years to more | 6,066 (70.09%) | 1,921 (63.51%) |
| **Educational Level** |  |  |
| No Level/Primary | 3,911 (42.11%) | 1,136 (34.20%) |
| Secondary/Superior | 5,375 (57.89%) | 2,186 (65.80%) |
| **Natural region** |  |  |
| Metropolitan Lima | 3,878 (32.35%) | 1,699 (42.25%) |
| Resy of coast | 3,289 (27.43%) | 1,163 (28.92%) |
| Mountain Range | 3,181 (26.53%) | 712 (17.71%) |
| Jungle | 1,640 (13.68%) | 447 (11.12%) |
| **Wealth index** |  |  |
| The poorest/Poor | 4,193 (34.98%) | 928 (23.07%) |
| Medium | 2,206 (18.40%) | 843 (20.96%) |
| Rich/Richest | 5,589 (46.62%) | 2,251 (55.97%) |
| **Area of residence** |  |  |
| Rural | 9,165 (76.45%) | 3,521 (87.53%) |
| Urban | 2,824 (23.55%) | 502 (12.47%) |
| **Nutritional Status** |  |  |
| Normal weight | 2958 (24.72%) | 939 (23.37%) |
| Overweight | 4730 (39.53%) | 1,587 (39.50%) |
| Obesity | 4276 (35.74%) | 1,492 (37.12%) |
| **Smoker Status** |  |  |
| Has never smoked | 10,620 (88.59%) | 3,520 (87.52%) |
| Former smoker | 500 (4.17%) | 177 (4.39%) |
| Currently smoker | 616 (5.14%) | 222 (5.53%) |
| Daily smoker | 251 (2.10%) | 103 (2.56%) |
| **Alcohol consumption** |  |  |
| Never or did not use in the last 12 months | 4,565 (38.08%) | 1,501 (37.32%) |
| Non-excessive | 7,153 (59.66%) | 2,412 (59.95%) |
| Excessive | 270 (2.26%) | 110 (2.73%) |
| **Fruit/vegetable consumption ≥ 5 servings per day** |  |  |
| No | 10,925 (91.13%) | 3,621 (90.02%) |
| Yes | 1,063 (8.87%) | 402 (9.98%) |
| **HTN diagnosis time** |  |  |
| < 2 years | 4,626 (38.59%) | — |
| ≥ 2 years | 7,362 (61.41%) | — |
| **Diabetes diagnosis time** |  |  |
| < 2 years | — | 1,217 (30.26%) |
| ≥ 2 years | — | 2,805 (69.74%) |

Among individuals diagnosed with hypertension, alcohol consumption patterns fluctuated significantly from 2014 to 2023. In 2014, 41.02% never consumed or did not consume in the last 12 months, 56.39% consumed moderately, and 2.59% excessively. The year 2020 marked a notable change, with an increase to 7.31% in excessive consumption. By 2023, moderate consumption increased to 58.32%, while abstinence decreased to 39.88%, and excessive consumption dropped to 1.80%. Regarding smoking, a general downward trend was observed from 2014 to 2023, with an increase in former smokers and a significant decrease in current and daily smokers in 2020. In 2023, the prevalence of current smokers remained stable, while daily smokers and former smokers have been slightly decreasing. Meanwhile, fruit and vegetable consumption reached its lowest point in 2020 (4.21%), increasing in 2023 without surpassing the maximum of the studied period. (Figure 1)

**Figure 1.**
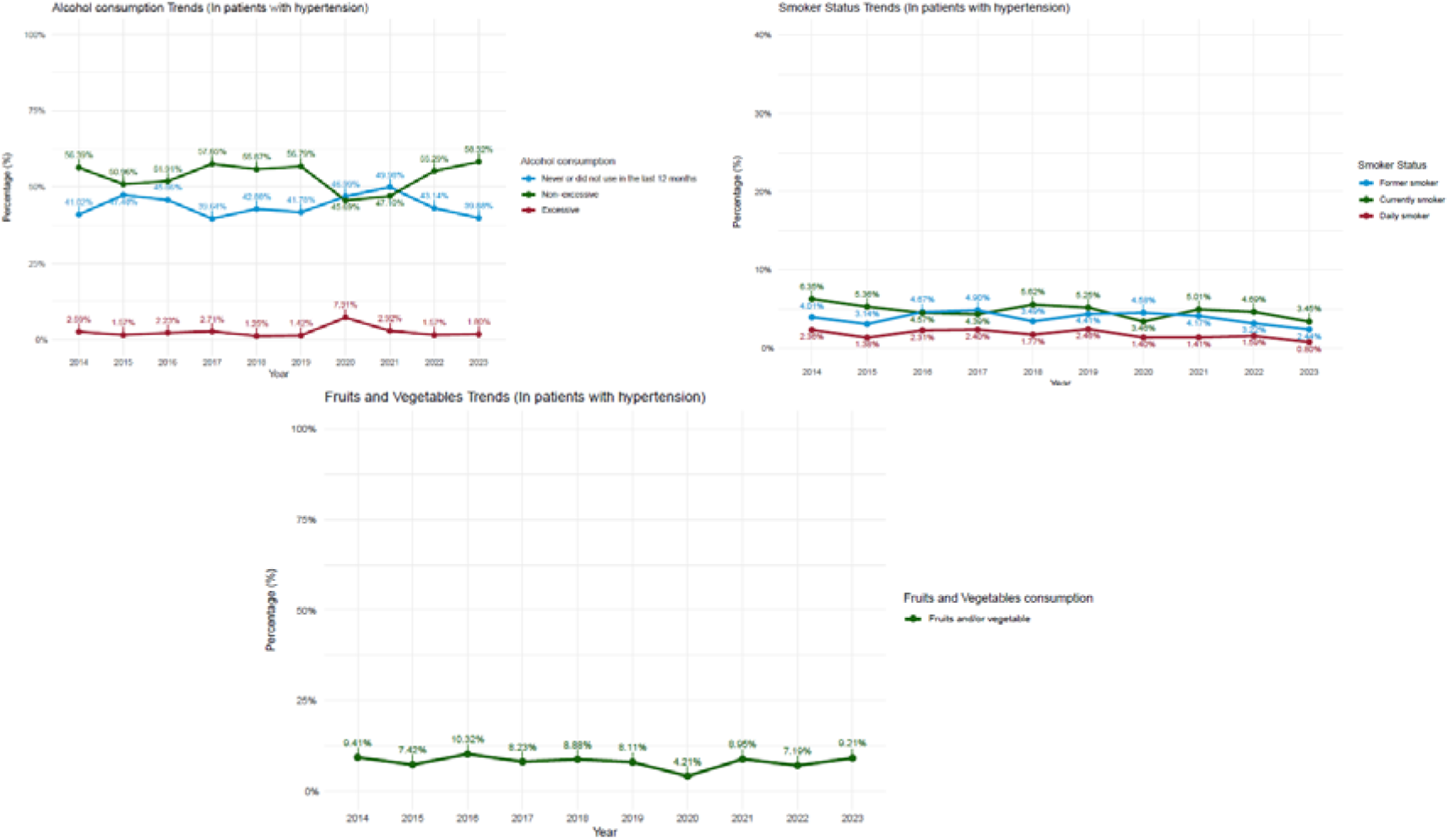
Chart of lifestyle trends in people diagnosed with high blood pressure between 2014 and 2023 according to the DHS.

In individuals with diabetes, alcohol consumption showed significant variations. In 2020, an alarming peak of 8.60% was observed in excessive consumption, while non-excessive consumption decreased to 51.59%. By 2023, these trends reversed: excessive consumption dropped to 1.55% and non-excessive consumption increased to 60.88%. Regarding smoking, 2017 presented similar prevalences across categories. Subsequently, current smokers increased, reaching their maximum in 2018, while daily smokers tended to decrease. In 2023, current smokers decreased by almost half compared to 2022, while daily smokers increased and former smokers reduced by a third. Meanwhile, fruit and vegetable consumption reached its lowest point (4.36%) during the studied period, followed by an increase to 11.93% the following year. However, it has not exceeded the maximum peak reached in 2018. (Figure 2)

**Figure 2.**
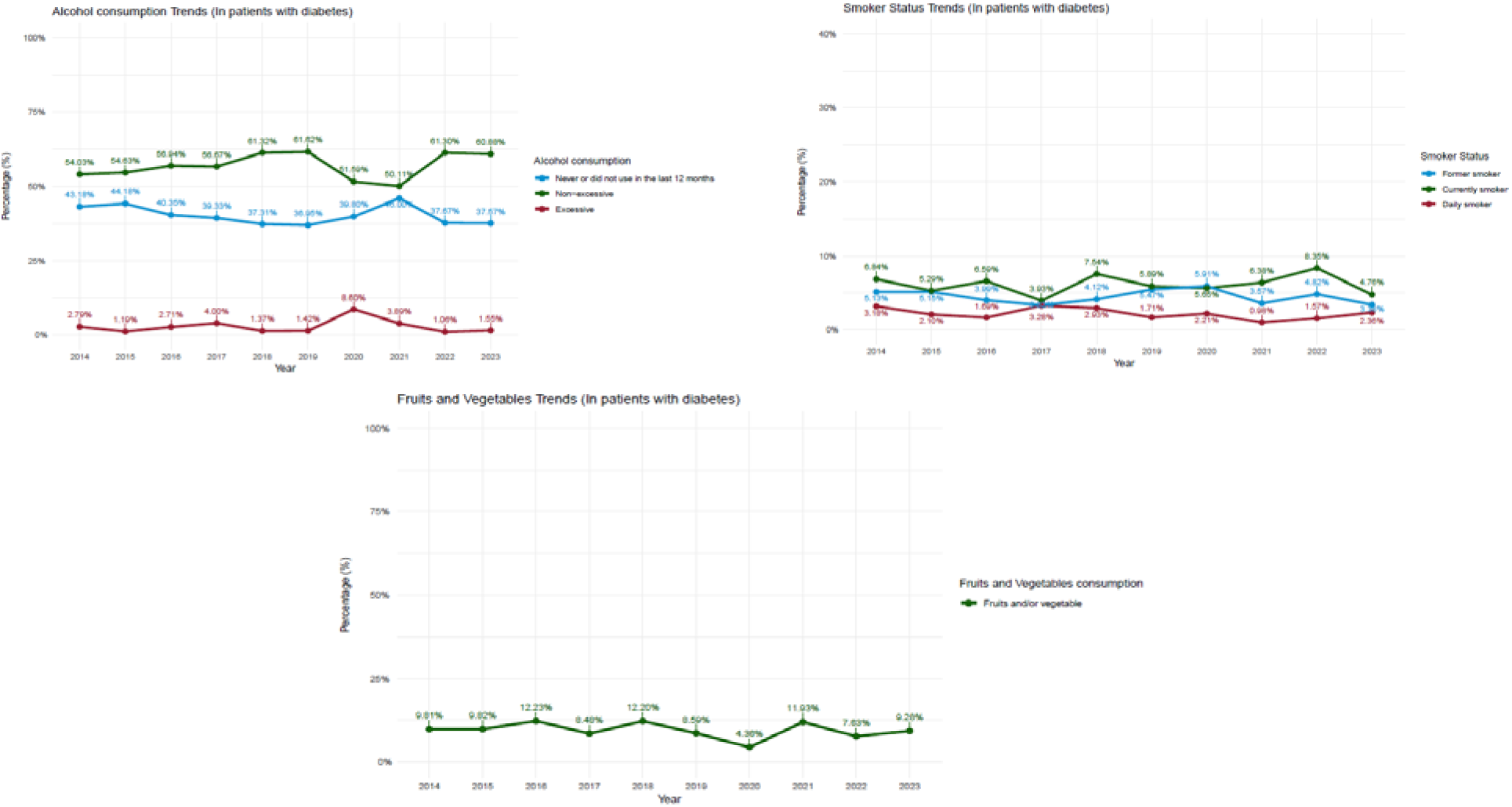
Chart of lifestyle trends in people diagnosed with diabetes between 2014 and 2023 according to the DHS

Table 2 shows that for patients with DM diagnosed for ≥ 2 years, the aPRs were 1.13 (95% CI: 0.83, 1.67) for smoking status, 0.90 (95% CI: 0.54, 1.56) for alcohol consumption, and 0.76 (95% CI: 0.59, 1.00) for fruit/vegetable consumption. In patients with HTN diagnosed for ≥ 2 years, the aPRs were 1.06 (95% CI: 0.86, 1.31), 0.79 (95% CI: 0.47, 1.38), and 1.77 (95% CI: 0.59, 1.01) respectively. These results suggest a statistically significant association between the time since diagnosis and the studied lifestyles, both in patients with DM and HTN, could not be established.

**Table 2.** Poisson regression analysis with robust variance between lifestyles and time since diagnosis of HTN and DM.

| Characteristic | Smoking Status | | Alcohol consumption | | Fruit/vegetable consumption $\geq 5$ servings | |
| --- | --- | --- | --- | --- | --- | --- |
|  | aPR* | 95% CI | aPR* | 95% CI | aPR* | 95% CI |
| **DM diagnosis time** |  |  |  |  |  |  |
| < 2 years | — | — | — | — | — | — |
| $\geq 2$ years | 1.13 | 0.83, 1.67 | 0.90 | 0.54, 1.56 | 0.76 | 0.59, 1.00 |
| **HTN diagnosis time** |  |  |  |  |  |  |
| < 2 years | — | — | — | — | — | — |
| $\geq 2$ years | 1.06 | 0.86, 1.31 | 0.79 | 0.47, 1.38 | 1.77 | 0.59, 1.01 |

Each factor has been independently adjusted for Sex, Group Age, Educational Level, Natural Region, Wealth Index, nutritional status, and area of residence
aPR: Prevalence ratio. CI 95%: Confidence interval at 95%
Source: self-made

## Discussion

### Prevalence and Trends of Lifestyles in Patients Diagnosed with Hypertension and Diabetes

Regarding alcohol consumption, trends revealed an increase in excessive alcohol consumption in both study groups in 2020, during the COVID-19 pandemic. These findings coincide with recent studies that have documented an increase in alcohol consumption among people with chronic diseases ^(12)^. This increase can be explained in different ways, including chronic stress associated with strategies for managing these diseases and the high-stress scenarios that the pandemic represented ^(13,14)^. Lack of social and emotional support can also increase dependence on alcohol as a coping mechanism for these personal sensations or emotions. In some cases, alcohol consumption may be related to depression, and there is a misconception that this substance can improve mood. At the same time, each person’s quality of life can contribute to excessive alcohol consumption ^(15,16)^.

Tobacco consumption patterns showed divergent trends between patients with hypertension and diabetes. In hypertensive patients, a general trend towards reduction was observed, with a notable increase in former smokers during the COVID-19 pandemic. In contrast, diabetics showed a trend towards an increase in daily smokers. Tobacco consumption in diabetics could be influenced by psychosocial factors and an underestimation of the associated risks, both of smoking habits and their interaction with diabetes. On the other hand, recent studies indicate that between 20% and 25% of hypertensive patients are smokers. The reduction of this habit during the COVID-19 pandemic in this group reflects not only a greater awareness of the general health risks of tobacco but also the fear of developing more severe forms of COVID-19, which motivated many to quit consumption. These findings underscore the complexity of factors influencing smoking habits in patients with chronic diseases and the importance of considering the global health context in health promotion strategies ^(17,18)^.

Tobacco use in people with chronic diseases such as diabetes and hypertension can be related to different factors such as patients’ living conditions, a social environment with prevalent tobacco habits, high stress levels, and low socioeconomic status ^(19)^. Additionally, in some cases, smoking may be associated with mental health comorbidity such as depression or anxiety, which are common in chronic patients. Patients could perceive tobacco consumption as a coping mechanism for the stress of emotional burdens related to living with chronic diseases, and fear of long-term disease complications can paradoxically lead some people to increase tobacco consumption in an attempt to alleviate their anxiety levels ^(20)^.

Finally, regarding fruit and vegetable consumption, the prevalence of this lifestyle dimension is very low in both study populations. These results are concerning because a diet rich in fruits and vegetables is essential for managing both conditions ^(21)^. A study conducted in Ecuador had very similar results to ours, as it found that in patients with risk factors for developing chronic non-communicable diseases, most participants had low fruit and vegetable consumption, preferring instead the consumption of industrialized foods such as sausages and canned goods, thus relating to unhealthy diet consumption ^(22,23)^. The low prevalence of consumption in our study population underscores the need for nutritional education programs and public policies that facilitate access to fresh fruits and vegetables.

One possible explanation for the low consumption of these foods could be the lack of adequate nutritional education; patients are not aware of the importance of these foods for managing these pathologies. Additionally, preferences and eating habits established over the years can be difficult to change due to cultural influences, and food traditions also play an important role as there are cultures that do not include a sufficient amount of fruits and vegetables in their daily diet. Another factor to consider is the perception of cost and effort involved in preparing these foods; some patients may perceive that good quality fruits and vegetables are more expensive or require more preparation time compared to processed foods. Finally, stress and lack of time can lead to quick and less healthy food choices ^(23,24)^.

### Association Between Lifestyles and Time Since Disease Diagnosis

The findings of the present study did not find a statistically significant association between the time since disease diagnosis and the presence of either comorbidity. This absence of variation in the lifestyles of these individuals could indicate that these habits remain stable and are not influenced by the duration of their diagnosis. This also suggests that interventions and programs aimed at improving lifestyles in patients with chronic diseases are not being effective in the long or short term in the study groups ^(26)^.

Although the time since diagnosis of a chronic disease could influence patients’ daily habits, behavior change is a complex process influenced by multiple factors. The perception of daily life plays a crucial role, as modifying ingrained lifestyles is challenging. Patients face various barriers to adopting healthier habits concerning alcohol consumption, tobacco use, and fruit and vegetable intake. These include limited access to reliable and understandable health information, scarcity of resources to maintain a healthy lifestyle (such as affordable nutritious foods or safe spaces for physical activity), and insufficient social support, which is fundamental for sustaining long-term changes. These obstacles underscore the need for a comprehensive approach to health promotion that not only considers the time since diagnosis but also addresses the socioeconomic and cultural barriers that hinder the adoption of healthy lifestyles^(27)^.

Furthermore, one reason why people with chronic diseases continue to consume alcohol, regardless of their diagnosis, is social and cultural influence. In many social settings, alcohol consumption is deeply entrenched insocial practices and is seen as an acceptable way to celebrate, relax, and socialize. It is precisely this social context that can make it difficult for patients to reduce or even eliminate their consumption, as it could imply a perception of exclusion within their social environment. Therefore, peer pressure and the influence of friends and family who also consume alcohol can play a significant role in the persistence of this habit, even in the presence of a chronic disease ^(28,29)^. Additionally, the lack of standardization in clinical guidelines for the diagnosis of hypertension and diabetes presents a significant challenge for medical practice and public health, as recommendations such as reducing alcohol consumption or consuming it in moderation may be subject to different interpretations, depending on who reads them ^(30,31)^. Thus, the variability in diagnostic criteria among different organizations can lead to inconsistencies in the diagnosis and treatment of these diseases.

The co-occurrence of other addictive behaviors and the permissive environment with tobacco or alcohol consumption should also be taken into account. In patients with chronic diseases, it has been shown that addiction alters brain systems related to reward and motivation, making quitting tobacco or alcohol truly challenging ^(32)^. From this concept, we can infer that people diagnosed with diabetes and hypertension, regardless of the duration of their illness, continue to consume alcohol and tobacco despite knowledge of the harmful effects of these substances on their pathologies. This could be influenced by risk perception and irrational optimism; patients may underestimate the severity of known consequences, believing that adverse effects will not directly affect them or that they can manage them. This irrational optimism can be reinforced by personal experiences or those of others who consume these substances without apparent immediate serious effects ^(33)^. Finally, it may also be related to a phenomenon called “sunk cost,” where people may perceive that they have invested time and resources in these habits, consequently considering it futile to abandon them at this point, believing that the damage has already been done.

Finally, the persistent low consumption of fruits and vegetables, regardless of the time since diagnosis, is a concerning finding but consistent with existing literature. Previous studies have shown that knowledge about the disease does not necessarily translate into better dietary practices. For example, Marcy et al. ^(34)^ found that only 12.4% of patients with diabetes consumed the recommended portions of fruits and vegetables, with no significant differences according to disease duration. Cost, availability, and cultural preferences are crucial in this behavior. Additionally, Nicklett and Kadell ^(35)^ pointed out that barriers to fruit and vegetable consumption in older adults with chronic diseases include physical limitations for food preparation and difficulties in accessing fresh produce. These findings underscore the need for multifaceted interventions that address nutritional education and socioeconomic and practical barriers to improve fruit and vegetable consumption in this population.

### Importance for Public Health

This study, based on DHS data in Peru, is of significant importance for public health. It presents a longitudinal analysis of lifestyles in people diagnosed with hypertension and diabetes, two chronic non-communicable diseases of high prevalence and considerable impact on population health.

The findings reveal complex patterns in the lifestyles of these populations. Notably, a decrease in smoking was observed among patients with hypertension, while this trend was not as marked in patients with diabetes. Alcohol consumption and fruit and vegetable intake showed fluctuations over time, without a clear trend towards sustained improvement.

The COVID-19 pandemic marked a turning point, causing significant changes in lifestyles, although many of these changes were temporary, and behaviors tended to return to levels similar to those before the pandemic. This observation underscores the complexity of modifying long-term habits and the need for sustained and adaptable intervention strategies.

The persistence of unhealthy lifestyles, especially low fruit and vegetable consumption, points to critical areas for nutritional interventions. This aspect is crucial, given that eating habits are closely related to the progression of diabetes and hypertension.

Thus, the results highlight the importance of continuous monitoring of health behaviors to identify trends and areas for improvement. Regular epidemiological surveillance can help policymakers develop more effective strategies, considering the observed differences between patients with hypertension and diabetes.

### Limitations of the Study

The design of this study was cross-sectional, which limits the causal approach of the study; however, the findings are considered important as a first step as an exploratory explanatory study. Another important limitation is that the data obtained for this study regarding the variables come from self-reports, which may lead to biases such as underestimation or overestimation of diabetes, hypertension, or the lifestyle dimensions taken into account in this research, in addition to memory biases on the part of the participants or a response related to social acceptance to the questions asked. Regarding the definitions of alcohol consumption, smoking status, and fruit and vegetable consumption, these may be variable and subject to interpretation, due to the lack of specific details such as quantities and frequencies of consumption of these elements. Due to the sample used, the presented results may not be extrapolated to different populations or contexts.

## Conclusions

Trends in lifestyles of people with hypertension and diabetes between 2014 and 2023 showed oscillations without sustained improvement. Although a decrease in daily smokers was observed in hypertensive patients in 2023, this was not seen in diabetics. Alcohol consumption and fruit and vegetable intake maintained fluctuating patterns without significant improvements. The time since diagnosis did not prove to be a determining factor for changes in lifestyles in both groups.

These findings suggest that prolonged coexistence with these chronic diseases does not automatically lead to the adoption of healthier habits, evidencing the need to implement more effective health education programs and strategies to promote healthy lifestyles in this population.

## Data Availability

The data supporting the findings of this study can be accessed by the original research paper at the follow link: https://proyectos.inei.gob.pe/microdatos/

## Acknowledgments

A special thanks to the members of Universidad Nacional Toribio Rodríguez de Mendoza de Amazonas (UNTRM), Amazonas, Peru for their support and contributions throughout the completion of this research.

## Informed consent

It was not necessary to obtain informed consent in this Study

## Authors’ contribution

Joan A. Loayza-Castro: Conceptualization, Investigation, Methodology, Resources, Writing - Original Draft, Writing - Review & Editing

Lupita Ana Maria Valladolid-Sandoval: Investigation, Validation, Visualization, Writing - Original Draft, Writing - Review & Editing

Luisa Erika Milagros Vásquez-Romero: Investigation, Project administration, Writing - Original Draft, Writing - Review & Editing

Fiorella E. Zuzunaga-Montoya: Investigation, Resources, Writing - Original Draft, Writing - Review & Editing

Jhonatan Roberto Astucuri Hidalgo: Software, Data Curation, Formal analysis, Writing - Review & Editing

Víctor Juan Vera-Ponce: Methodology, Supervision, Funding acquisition, Writing - Review & Editing

